# Genome-wide association study across five cohorts identifies five novel loci associated with idiopathic pulmonary fibrosis

**DOI:** 10.1101/2021.12.06.21266509

**Authors:** Richard J Allen, Amy Stockwell, Justin M Oldham, Beatriz Guillen-Guio, Carlos Flores, Imre Noth, Brian L Yaspan, R Gisli Jenkins, Louise V Wain, International IPF Genetics Consortium

## Abstract

Idiopathic pulmonary fibrosis (IPF) is a chronic lung condition with poor survival times. We previously published a genome-wide meta-analysis of IPF risk across three studies with independent replication of associated variants in two additional studies. To maximise power and to generate more accurate effect size estimates, we performed a genome-wide meta-analysis across all five studies included in the previous IPF risk GWAS. We utilised the distribution of effect sizes across the five studies to assess the replicability of the results and identified five robust novel genetic association signals implicating mTOR signalling, telomere maintenance and spindle assembly genes in IPF risk.

## Introduction

Idiopathic pulmonary fibrosis (IPF) is a chronic lung disease believed to result from an aberrant response to alveolar injury leading to a build-up of scar tissue. This progressive scarring is eventually fatal with half of individuals dying within 3 to 5 years of diagnosis^1^. The cause of IPF is unknown but genetics play an important role in how susceptible an individual is to IPF^2^.

Genome-wide association studies (GWAS) are an approach whereby genetic variants from across the genome are tested for their association with a disease. Genetic loci identified by GWAS can implicate genes important in disease pathogenesis and drugs which target the products encoded by these genetically-supported genes are twice as likely to be successful during development. The genetic association statistics from a GWAS are also widely used to identify causal markers of disease through Mendelian randomisation, to conduct heritability estimation and for genetic correlation analyses. It is therefore important that sample sizes are maximised to ensure sufficient statistical power to detect genetic associations and to generate precise effect size estimates.

We recently published a GWAS of IPF risk^2^. The discovery GWAS consisted of three studies (named as the UK, Chicago and Colorado studies) and a replication analysis performed in two independent studies (named as the UUS [USA, UK and Spain] and Genentech studies). This analysis reported 14 genetic signals which implicated host defence, cell-cell adhesion, spindle assembly, TGF-β signalling regulation and telomere maintenance as important biological processes involved in IPF disease risk.

We here present a meta-analysis of genome-wide data from all 5 datasets included in our previous study. The results of this analysis implicate new genetic loci in IPF pathogenesis and provide a unique resource for other studies of IPF risk and pathogenesis.

## Methods

Quality control and sample selection have been previously described^2^. In summary, all datasets comprised of unrelated European-ancestry individuals. Individuals in the Genentech study were sequenced using HiSeq X Ten platform (Illumina) and all other individuals were imputed from genotyping data using the HRC reference panel^3^. Genome-wide analyses were performed in each study separately using an additive logistic regression model adjusting for the first 10 genetic principal components to account for population stratification.

The individual GWAS results from the five studies were meta-analysed using an inverse-variance weighted fixed effect meta-analysis using METAL^4^. Variants were included in the meta-analysis if they were available in at least four studies. Genomic control was performed on the meta-analysis results using the LD score regression intercept to account for inflation not explained by polygenic effects^5^. Significant variants were defined as those with meta-analysis p<5×10^−8^ and conditional analyses were performed using GCTA-COJO to identify additional independent associated variants^6^. Independent associated variants were defined as variants remaining genome-wide significant after conditioning on the most significant variant (sentinel) in the region with consistent effect size estimates in the conditional and non-conditional analysis. Annotation of the sentinel variants was then performed using Variant Effect Predictor^7^.

To assess the robustness of novel results, we tested the strength and consistency of results across studies using MAMBA (Meta-Analysis Model-based Assessment of replicability)^8^. Variants with a posterior probability of replicability (PPR)≥90% were considered robust and likely to replicate should additional independent datasets become available.

Genome-wide summary statistics can be accessed at https://github.com/genomicsITER/PFgenetics.

## Results

A total of 4,125 cases, 20,464 controls and 7,554,248 genetic variants were included in the analysis (**Figure 1**). The UUS study included one additional case (due to resolving a sample ID issue since the previous publication) and one fewer control (where the individual has since withdrawn consent from UK Biobank) than described in the previous GWAS^2^.

**Figure 1:**
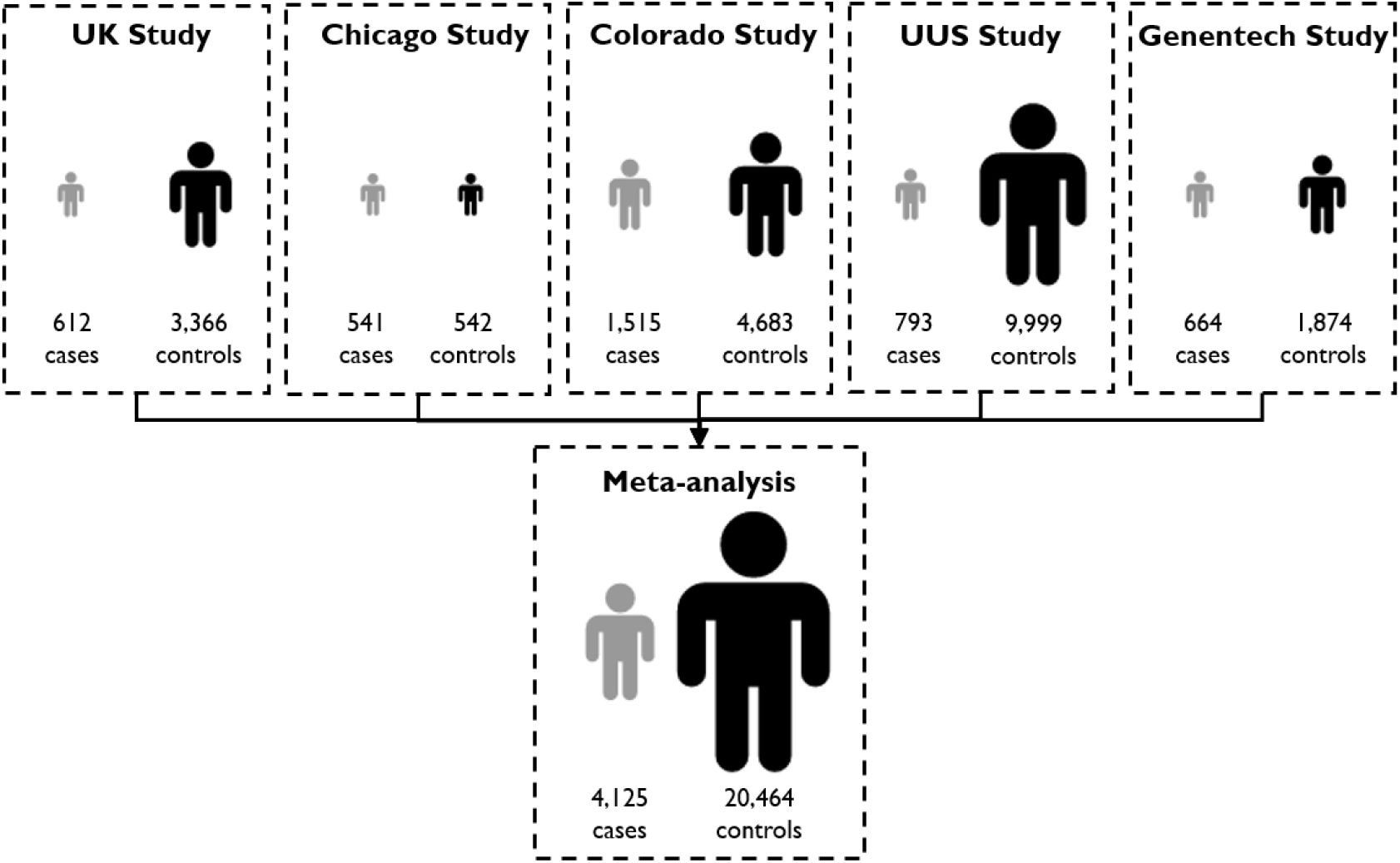
Study design and sample sizes.

After conditional analyses, there were 23 independent signals with p<5×10^−8^ in the genome-wide meta-analysis (**Figure 2**). These 23 signals included all 14 associations reported in the previous GWAS (**Supplementary Table 1**). Of the nine novel genetic associations (**Table 1**), five showed evidence of replicability (PPR≥90%). The sentinel variants of these five loci included variants in introns of *KNL1, NPRL3, STMN3* and *RTEL1*, and an intergenic variant in 10q25.1. All five novel variants had consistent direction of effect across all of the individual studies and reached nominal significance (p<0.05) in at least 3 of the studies.

**Figure 2:**
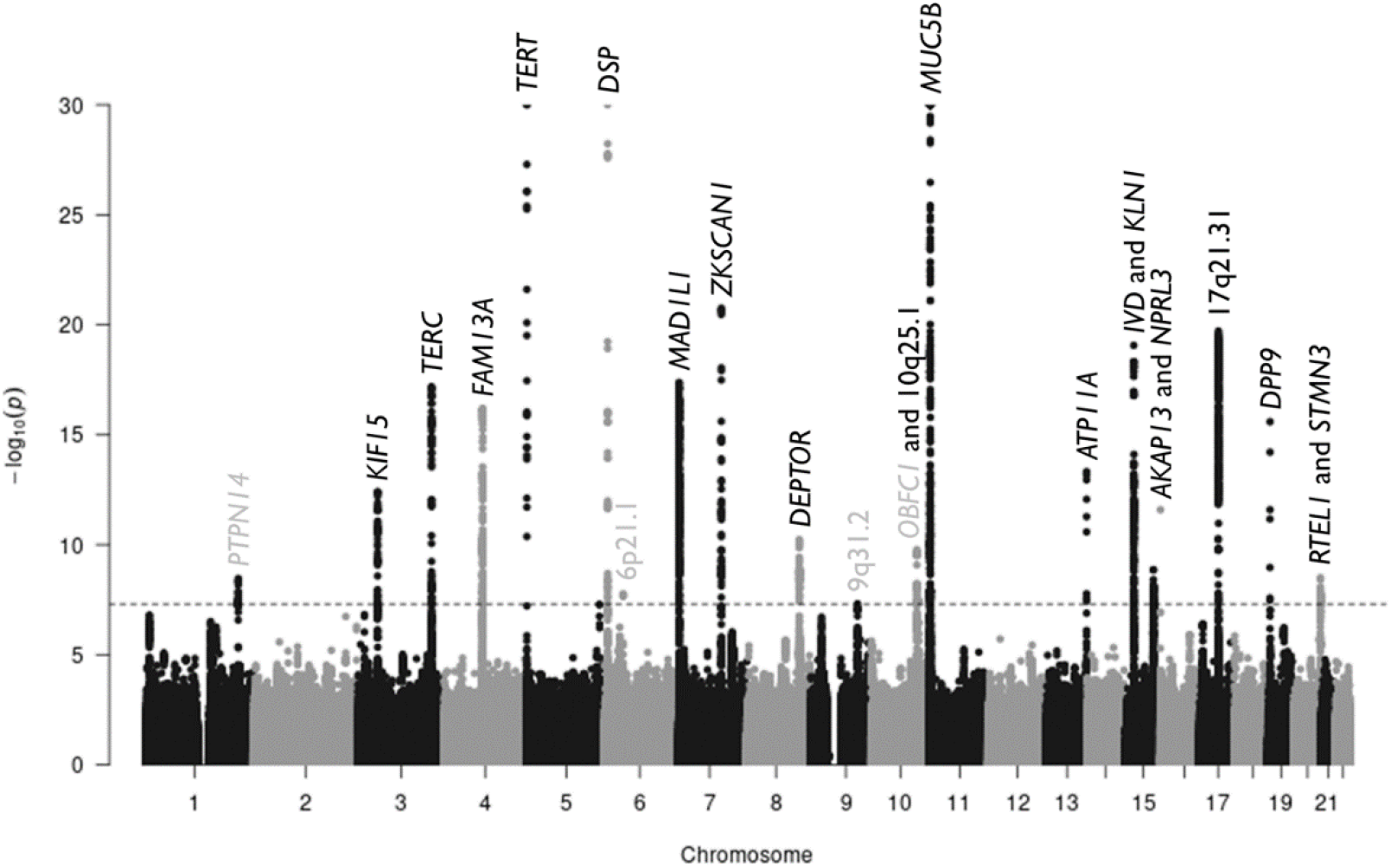
Manhattan plot. Each point shows a genetic variant with chromosomal position on the x axis and the –log(p value) on the y axis. The grey dashed line shows the genome-wide significance level (p=5×10^−8^). Each signal is labelled with the gene implicated by that signal. Genes in grey are the novel loci that do show evidence of replicability. The plot has been truncated at p=10^−30^.

**Table 1:**
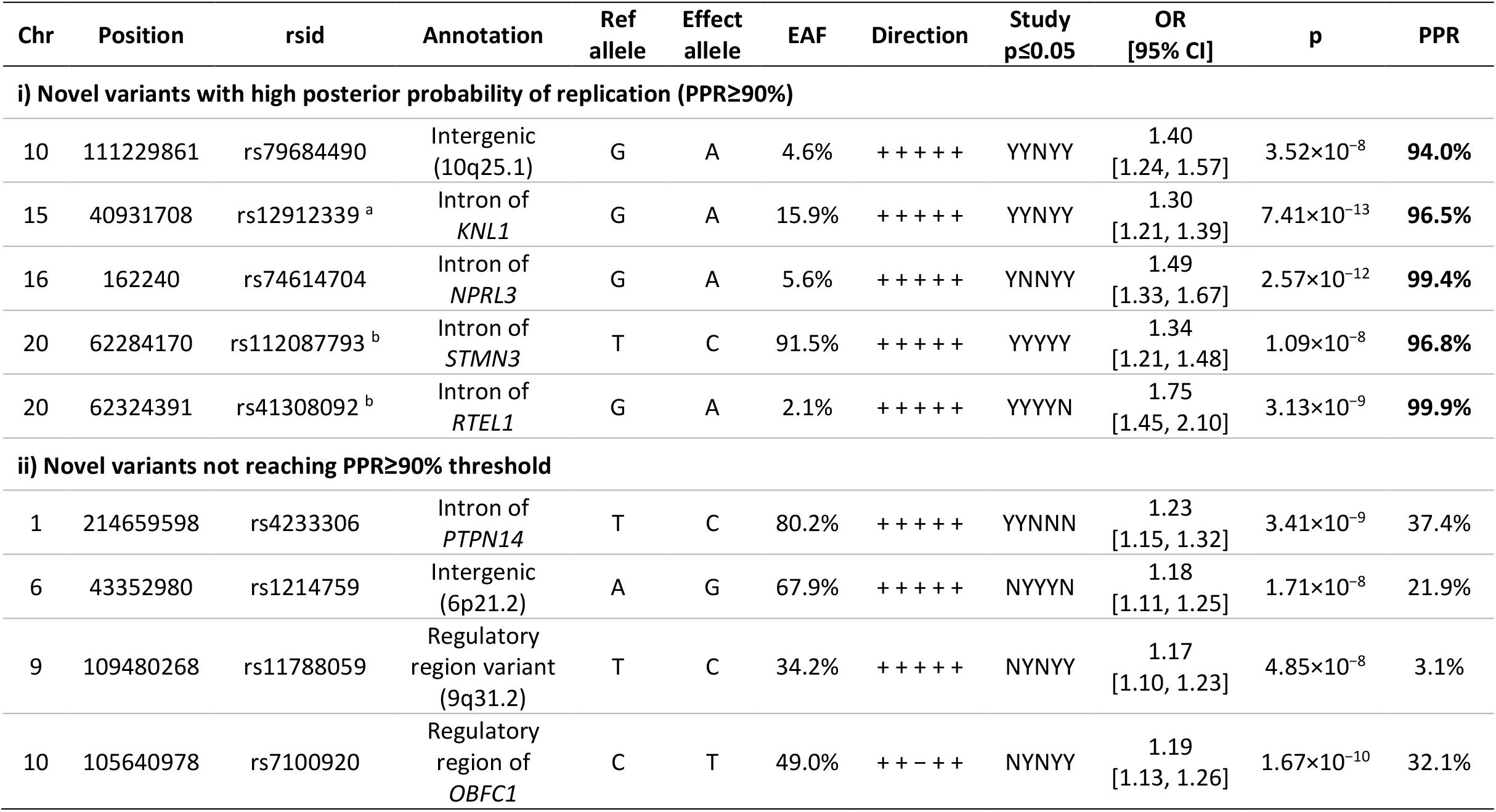
Sentinel variants of novel associations. Novel variants are defined as those not reaching significance criteria in previous analysis^2^ (the *RTEL1* and *OBFC1* signals have previously shown a possible association – see discussion). Effect sizes and directions are given in terms of the allele that increases risk of IPF. Chr=Chromosome. Position is based on genetic build 37. Annotation obtained from Variant Effect Predictor^7^. EAF=Effect allele frequency calculated across the five studies. The “Direction” column shows the direction of the beta in each of the five individual studies (+ means beta>0, − means beta<0). The “Study p≤0.05” column denotes which individual studies the variant reached nominal significance in (Y means p≤0.05, N means p>0.05). Both the direction and study p<0.05 are given in the order UK, Colorado, Chicago, UUS and then Genentech. OR=Odds ratio. CI=Confidence interval. PPR=posterior probability of replicability calculated using MAMBA^8. a^ The signal at *KNL1* is independent of the previously reported nearby signal in the *IVD* gene. ^b^ The *RTEL1* and *STMN3* signals are independent of each other.

## Discussion

By increasing the number of cases in the discovery analysis by more than 50% compared with the previous IPF risk GWAS, we identified novel genetic signals associated with IPF risk and improved the precision of estimations for previously reported signals. The five novel loci had internal evidence of replicability giving us confidence that these signals are likely to be generalisable.

The signals in *RTEL1* and *OBFC1* have been reported previously but did not meet the significance criteria of the previous three-way GWAS^2^. The new MAMBA analysis suggests that the consistency of effect across studies provides high confidence that the *RTEL1* signal will replicate should an independent dataset become available. This is not the case for the *OBFC1* signal where a low posterior probability of replication suggests that there may be heterogeneity in effect across the contributing studies.

The novel signals require further characterisation to determine the likely causal gene and underlying functional effect of the variants. However, some of the genes that are closest to these new signals have strong candidacy for involvement in IPF pathogenesis. *NPRL3* encodes a GATOR1 complex function component and acts through mTORC1 signalling to inhibit mTOR kinase activity^9^. mTOR regulates TGF-β collagen synthesis and inhibiting mTOR leads to increased deposition of scar tissue^10^. We previously reported an association implicating *DEPTOR*, another mTOR inhibiting gene. We also add to the evidence that cellular ageing plays a key role in IPF pathogenesis through associations at the telomere maintenance genes *TERT, TERC* and *RTEL1*. We previously reported associations in spindle assembly genes (*MAD1L1* and *KIF15*) and have identified a novel genetic association in another spindle assembly gene *KNL1* (Kinetochore Scaffold 1 also known as *CASC5*). *STMN3* (Stathmin 3) implicates another cell replication process through tubulin binding^9^.

By maximising the statistical power of the analysis, we identified novel genetic associations with IPF risk. These signals may implicate biologically relevant genes that support the importance of TGF-β signalling and cell replication as important processes in disease pathogenesis.

## Data Availability

Genome-wide summary statistics can be accessed at https://github.com/genomicsITER/PFgenetics.

## Ethics Statement

This research was conducted using previously published work with appropriate ethics approval. The PROFILE study (which provided samples for the UK and UUS studies) had institutional ethics approval at the University of Nottingham (NCT01134822 – ethics reference 10/H0402/2) and Royal Brompton and Harefield NHS Foundation Trust (NCT01110694 – ethics reference 10/H0720/12). Spanish samples were recruited under ethics approval by ethics committee from the Hospital Universitario N.S. de Candelaria (reference of the approval: PI-19/12). The UUS study also included individuals from clinical trials with ethics approval (ACE [NCT00957242] and PANTHER [NCT00650091]). UK samples were recruited across multiple sites with individual ethics approval (University of Edinburgh Research Ethics Committee [The Edinburgh Lung Fibrosis Molecular Endotyping (ELFMEN) Study NCT04016181] 17/ES/0075, NRES Committee South West – Southmead, Yorkshire and Humber Research Ethics Committee 08/H1304/54 and Nottingham Research Ethics Committee 09/H0403/59). For individuals recruited at the University of Chicago, consenting patients with IPF who were prospectively enrolled in the institutional review board-approved ILD registry (IRB#14163A) were included. Individuals recruited at the University of Pittsburgh Medical Centre had ethics approval from the University of Pittsburgh Human Research Protection Office (reference STUDY20030223: Genetic Polymorphisms in IPF). Individuals from the COMET (NCT01071707) and Lung Tissue Research Consortium (NCT02988388) studies were also included in the Chicago study. All subjects in the Colorado study gave written informed consent as part of IRB-approved protocols for their recruitment at each site and the GWAS study was approved by the National Jewish Health IRB and Colorado Combined Institutional Review Boards (COMIRB). Subjects in the Genentech study provided written informed consent for whole-genome sequencing of their DNA. Ethical approval was provided as per the original clinical trials (INSPIRE [NCT00075998], RIFF [NCT01872689], CAPACITY [NCT00287729 and NCT00287716] and ASCEND [NCT01366209]). For the USCF cohort, sample and data collection were approved by the University of California San Francisco Committee on Human Research and all patients provided written informed consent. For the Vanderbilt cohort, the Institutional Review Boards from Vanderbilt University approved the study and all participants provided written informed consent before enrolment.

## Supplement

### International IPF Genetics Consortium

#### Writing Group

Richard J Allen, Carlos Flores, Beatriz Guillen-Guio, R Gisli Jenkins, Imre Noth, Justin M Oldham, Amy Stockwell, Louise V Wain, Brian L Yaspan

#### UK Study

Richard J Allen, Helen L Booth, William A Fahy, Ian P Hall, Simon P Hart, Mike R Hill, Nik Hirani, Richard B Hubbard, R Gisli Jenkins, Toby M Maher, Robin J McAnulty, Ann B Millar, Philip L Molyneaux, Vidya Navaratnam, Eunice Oballa, Helen Parfrey, Gauri Saini, Ian Sayers, Martin D Tobin, Louise V Wain, Moira K B Whyte

#### Chicago Study

Ayodeji Adegunsoye, Carlos Flores, Naftali Kaminski, Shwu-Fan Ma, Imre Noth, Justin M Oldham, Mary E Strek, Yingze Zhang

#### Colorado Study

Tasha E Fingerlin, David A Schwartz

#### UUS Study

Richard J Allen, Carlos Flores, Beatriz Guillen-Guio, R Gisli Jenkins, Shwu-Fan Ma, Toby M Maher, Maria Molina-Molina, Philip L Molyneaux, Imre Noth, Justin M Oldham, Louise V Wain

#### Genentech study

Margaret Neighbors, Xuting Sheng, Amy Stockwell, Brian L Yaspan

**Supplementary Table 1:** All sentinel variants associated with IPF risk. This table includes the most associated variant (sentinel) for the 19 signals (14 previously reported loci and the five novel loci identified here) associated with IPF risk. The risk allele is the allele associated with increased risk of IPF. Position is for genetic build 37. Chr=chromosome. EAF=Effect allele frequency. OR=Odds ratio. CI=Confidence interval.

| Chr | Position | rsid | Implicated gene | Non-effect allele | Risk allele | EAF | OR [95% CI] | p |
| --- | --- | --- | --- | --- | --- | --- | --- | --- |
| 3 | 44903434 | rs2292181 | *KIF15* | G | C | 5.2% | 1.52<br>[1.36, 1.70] | 3.95×10<sup>-13</sup> |
| 3 | 169486271 | rs9860874 | *TERC* | C | A | 27.6% | 1.29<br>[1.22, 1.37] | 6.49×10<sup>-18</sup> |
| 4 | 89837808 | rs2609259 | *FAM13A* | C | A | 22.4% | 1.30<br>[1.22, 1.39] | 6.47×10<sup>-17</sup> |
| 5 | 1282414 | rs7725218 | *TERT* | A | G | 67.1% | 1.41<br>[1.33, 1.50] | 4.90×10<sup>-32</sup> |
| 6 | 7563232 | rs2076295 | *DSP* | T | G | 46.7% | 1.49<br>[1.41, 1.57] | 1.50×10<sup>-48</sup> |
| 7 | 1868761 | rs12537430 | *MAD1L1* | A | G | 62.5% | 1.28<br>[1.21, 1.35] | 4.20×10<sup>-18</sup> |
| 7 | 99630342 | rs2897075 | *ZKSCAN1* | C | T | 38.2% | 1.30<br>[1.23, 1.37] | 1.77×10<sup>-21</sup> |
| 8 | 120940206 | rs10808505 | *DEPTOR* | G | T | 57.3% | 1.20<br>[1.13, 1.26] | 6.03×10<sup>-11</sup> |
| 10 | 111229861 | rs79684490 | 10q25.1 | G | A | 4.6% | 1.40<br>[1.24, 1.57] | 3.52×10<sup>-8</sup> |
| 11 | 1241221 | rs35705950 | *MUC5B* | G | T | 14.5% | 5.06<br>[4.69, 5.47] | 9.09×10<sup>-418</sup> |
| 13 | 113534984 | rs9577395 | *ATP11A* | G | C | 79.1% | 1.29<br>[1.21, 1.38] | 4.78×10<sup>-14</sup> |
| 15 | 40716253 | rs2304645 | *IVD* | G | C | 52.6% | 1.28<br>[1.21, 1.35] | 8.66×10<sup>-20</sup> |
| 15 | 40931708 | rs12912339 | *KNL1* | G | A | 15.9% | 1.30<br>[1.21, 1.39] | 7.41×10<sup>-13</sup> |
| 15 | 86287910 | rs11073517 | *AKAP13* | C | T | 32.7% | 1.19<br>[1.13, 1.26] | 1.36×10<sup>-9</sup> |
| 16 | 162240 | rs74614704 | *NPRL3* | G | A | 5.6% | 1.49<br>[1.33, 1.67] | 2.57×10<sup>-12</sup> |
| 17 | 44214888 | rs2077551 | 17q21.31 | C | T | 80.7% | 1.42<br>[1.32, 1.53] | 1.92×10<sup>-20</sup> |
| 19 | 4717672 | rs12610495 | *DPP9* | A | G | 30.6% | 1.28<br>[1.21, 1.36] | 2.58×10<sup>-16</sup> |
| 20 | 62284170 | rs112087793 | *STMN3* | T | C | 91.5% | 1.34<br>[1.21, 1.48] | 1.09×10<sup>-8</sup> |
| 20 | 62324391 | rs41308092 | *RTEL1* | G | A | 2.1% | 1.75<br>[1.45, 2.10] | 3.13×10<sup>-9</sup> |

